# THE USE OF COMPLEMENTARY PRACTICES BY PATIENTS UNDERGOING MEDICALLY ASSISTED REPRODUCTION: The place of Chinese medicine and related practices

**DOI:** 10.1101/2024.07.26.24311042

**Authors:** O. Serri, A. Meunier, P.E. Bouet, H. El Hachem, B. Liu, P. May-Panloup

## Abstract

**Purpose:** Infertility is associated with different factors, highly intricated but sometimes ill-defined. Despite the significant recent technological and therapeutic advancements, the success rates of assisted reproductive technologies (ART) remain relatively low. In this context several women have recourse to complementary practices (CP), in the hope of improving success rates and lowering the associated strain. These methods, are quite heterogenous, and are yet to be comprehensively analyzed in the literature.

The objective of our study was to evaluate, in women consulting at our fertility center, the current interest in these CP and more specifically, the role of Traditional Chinese Medicine (TCM) and associated Mind Body Intervention (MBI).

**Methods:** A prospective monocentric survey study at the fertility center of the Angers University Hospital between January and May 2022.

**Results:** 150 questionnaires were analyzed. Approximately 50% of women have recourse to CP, in the hope of improving the success rates of ART and reducing the associated stress and fatigue. The main CP used were the ones based on body manipulation, TCM and MBI. The lack of use was mainly linked to a lack of knowledge of these methods, and the vast majority of women would like to receive additional information about CP from the physicians at the center.

**Conclusion:** Our study shows a significant interest in CP in reproductive medicine. In view of the benefits of some of these practices, at least on the quality of life of patients, information on their value should be made available to all couples undergoing ART.

## INTRODUCTION

Infertility affects one in ten couples and, in France, approximately 3% of live births are the result of assisted reproductive technologies (ART) (1). However, and despite the recent technological advancements, success rates remain relatively low, with live birth rates being 10.3% per intrauterine insemination (IUI) cycle, and 18.5% per *in vitro* fertilization (IVF) cycle (1).

The diagnosis and treatment of infertility can cause significant psychosocial distress for the couple, which can manifest as emotional disturbances (2) and could, in return, negatively impact the chances of success of ART (3). The difficulty of dealing with infertility, coupled with the long and arduous process of IVF, and the lack of guaranteed results, can sometimes push couples to seek complementary methods of treatment, with different motives and expectations. In recent years, the use of these complementary methods, also referred to as “add-ons”, has significantly grew in the field of infertility and ART (4,5). They comprise of procedures, techniques, and medications that are used as complements to the standard ART protocols.

The first category includes the use of variants of the clinical and biological techniques used in classical protocols. These have been analyzed and listed by the HEFA in order to provide all patients easy and accessible information based on evidence-based medicine (6).

The second category includes the complementary practices (CP) used by patients, most often at their own initiative, despite the lack of any exhaustive analysis of these techniques, their mechanisms of action, objectives or efficiency. These treatments are highly popular nowadays, with a 2018 HEFA survey revealing that >50% of patients used one of them (acupuncture (26%), massages (11%), meditation (9%), and others (7%))(7).

Traditional Chinese medicine (TCM) is a medical system that is more than 3000 years old. Despite being considered a complimentary therapy in Western countries, it is the primary medicine in several Asian countries. It includes acupuncture, traditional pharmacopeia, and -mind-body interventions (MBI) (including manipulation techniques such as Tuina massage). Its efficiency in the field of infertility could rely on the adjustment of the hypothalamic-pituitary axis, the improvement of the ovarian function and the oocyte quality (especially in cases of decreased ovarian reserve), the enhancement of endometrial receptivity (vascularization and expression of hormone receptors), and the reduction of several inflammatory processes inherent to certain types of infertility (8).

The primary objective of our study was to evaluate, in women consulting at our fertility center for infertility, the interest in these complementary methods, and the reasons behind it. More precisely, our aim was to assess, in this patient population, the knowledge of TCM and related MBI, as well as the rate of recourse.

## MATERIALS AND METHODS

### Study protocol

We performed a prospective, monocentric descriptive and analytic study, with the purpose of describing and analyzing the complementary medical practices spontaneously used by women being managed for infertility. The survey was proposed to all women presenting to our fertility center, whether for an infertility work-up, fertility treatment (intrauterine insemination (IUI), IVF with or without ICSI), or for a post treatment follow-up.

### Methods

An information letter and an anonymous survey on complementary and alternative practices in ART were printed and made available at the fertility center of the Angers University Hospital. The secretaries of the department offered them to all women presenting to the center between January and May 2022. Women who anonymously answered the survey put the papers back in a dedicated container in the waiting room. The container was regularly emptied, and the surveys collected by our research fellow.

We performed a descriptive analysis of the completed surveys: the complementary practices most used by patients, the sources of information that led to their use, and the motivation behind their pursuit. We compared the rate of recourse to complementary practices according to the patients’ age, the education level, and the reason for their care in the center. The parametric Student’s t-test was used for quantitative variables, and the chi-squared (χ2) test was used for qualitative variables. All statistical tests were considered significant at p-value < 0.05.

- ***Inclusion criteria*** All patients who presented to the fertility unit and answered the survey.
- ***Non-inclusion criteria***

All patients who refused to complete the survey.

### Ethical and regulatory aspects

The study received a favorable opinion from the Ethics committee of the Angers University Hospital, registered under the number 2021-213. The study is registered at the “Commission Nationale de l’informatique et des libertés” (CNIL) under the number ar21-0162v0.

## RESULTS

### Population characteristics

In total, 150 women completed the survey.

-Mean age of patients was 32.9 5.2 (± 5.2) years.

-The causes of consultation at the fertility unit were: 44% for an IVF/ICSI treatment, 27% for an infertility work-up, 7% for an IUI, 7% for controlled ovarian stimulation (COS) followed by intercourse, 3% for post treatment follow-up, and 11% for other reasons (oocyte cryopreservation…)

Concerning the education level, 41% of participating women had a license degree, 26% had a Bachelor’s degree, 22% had a Master’s degree, and 11% had a high school degree.

### Patients using CP and their main reasons

In our center 48.7% of the 150 women who filled the survey have recourse to CP (figure 1 A) These patients mainly use CP in order to: Improve the success rate of ART, lower stress and fatigue levels.

**Figure 1:**
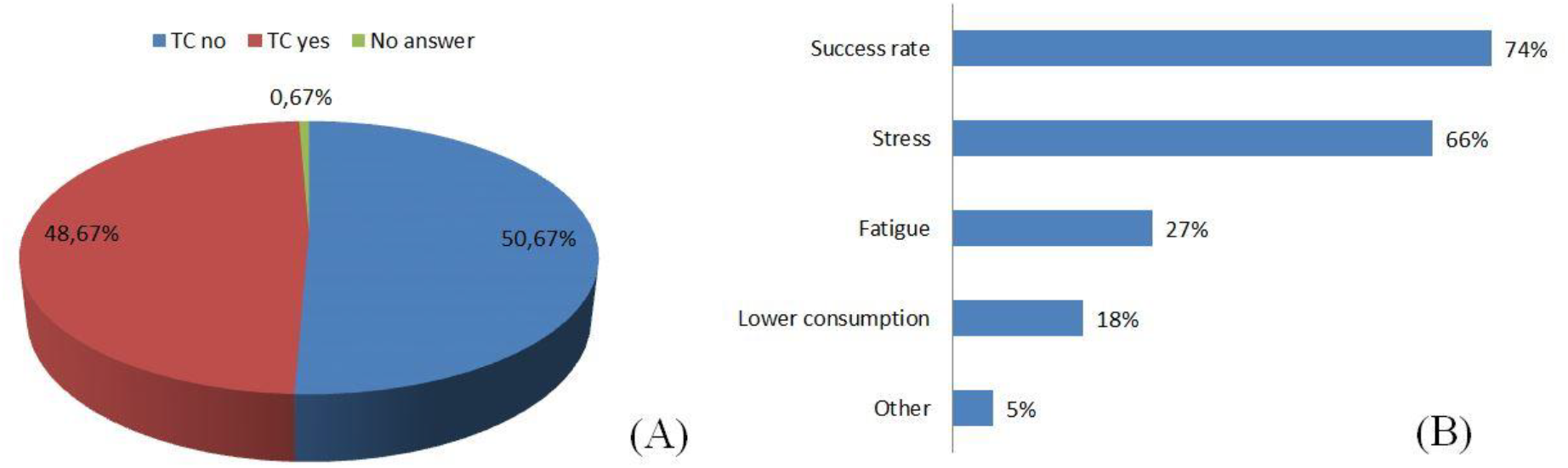
Sector diagram showing CP use in our population. In red the percentage of patients using CP, in blue the percentage of patients not using them and in green the percentage of non-response **(A)**. Bar chart showing the main reasons why patients use CP, Lower consumption = help reduce the consumption of substances that might negatively impact fertility (Tobacco…) **(B)**.

### Reason for the care in the center according to the use or no of CP

There was a significant difference (p=0.001) in the type of fertility treatment between women who used and those who did not use CP: The majority of patients using CP were undergoing IVF/ICSI, while the majority of women not using CP were undergoing infertility workup.

### Use of CP according to the educational level

There was no difference in the rate of recourse to CP according to the educational level (p=0.1)

### Use of CP according to age

There was no age difference between women who used and those who did not use CP (32.9 vs 32.8 years, p =0.9, resepctively)

## Type of CP used

The most commonly used CP are : Techniques based on body manipulation (O/C/M : Ostheopathy/chiropractic/massage), Mind-Body interventions (MBI), and traditional chinese medicine (TCM)

### Details of Mind-Body Interventions (MBI) (n=38) *(Figure 5)*

Among the MBI, we noted a large heterogeneity of methods. None of the patients used Qi-Gong (QG) or Tai-Chi (Tchi).

### Mode of Discovery of CP

The personal network was the most common mode of dissemination of CP. Healthcare professionals (moslty midwives) play an important role.

### Reasons for not using CP and desire for information

The majority of patients who did not use CP underline their lack of knowledge of these methods.

Almost all patients who participated would like to receive information on CP from the physicians at the fertility unit. Most frequently, they want the information to be based on scientific proof, but a large part wants exhaustive information

## DISCUSSION

Complementary and alternative medicine is defined by the World Health Organization (WHO) as a “broad set of health care practices that are not part of that country’s own tradition or conventional medicine and are not fully integrated into the dominant health care system” (9). The WHO counts more than 400 complementary and alternatives medicines, including methods based on natural products (phytotherapy, aromatherapy), techniques based on manipulation (osteopathy, chiropractic…), mind-body interventions (hypnotherapy, meditation, sophrology…), or complete systems based on specific theories (Traditional Chinese Medicine, homeopathy…). These practices are quite heterogeneous, and it is impossible to discuss them as a single homogeneous group.

The use of these complementary practices is widespread in certain medical fields, most notably oncology. However, in the field of infertility, and more specifically during ART, their use is yet to be fully analyzed, on a French national or international level.

Infertility can be caused by chronic pathologies with mainly biologic determinants, but also environmental and psychosomatic linked to an important emotional charge in patients. CP offer a different form of accompaniment for these patients during their infertility journey, by offering support, helping them relax, and making them feel capable of maximizing their chances of success (10).

Our study has shown that a considerable number of women (48.7%) (Figure 1) have recourse to CP, a finding concordant with the literature. Indeed, the HEFA 2018 study had shown a 50% rate of recourse to CP among women treated for infertility (11). Likewise, an Australian study reported a 66% rate (12) while an American study of 428 couples reported a 29% rate (13).

The main purposes, behind the recourse to CP, were to improve the pregnancy rates, and reduce the stress and fatigue associated with infertility treatments (Figure 2). It is worth noting that the utilization rate was higher among women undergoing IVF-ICSI treatment, which is widely known to be a long and arduous process (Figure 3). The most commonly employed CP were the ones based on body manipulation, mind-body interventions, and TCM (especially acupuncture) (Figure 4).

**Figure 2:**
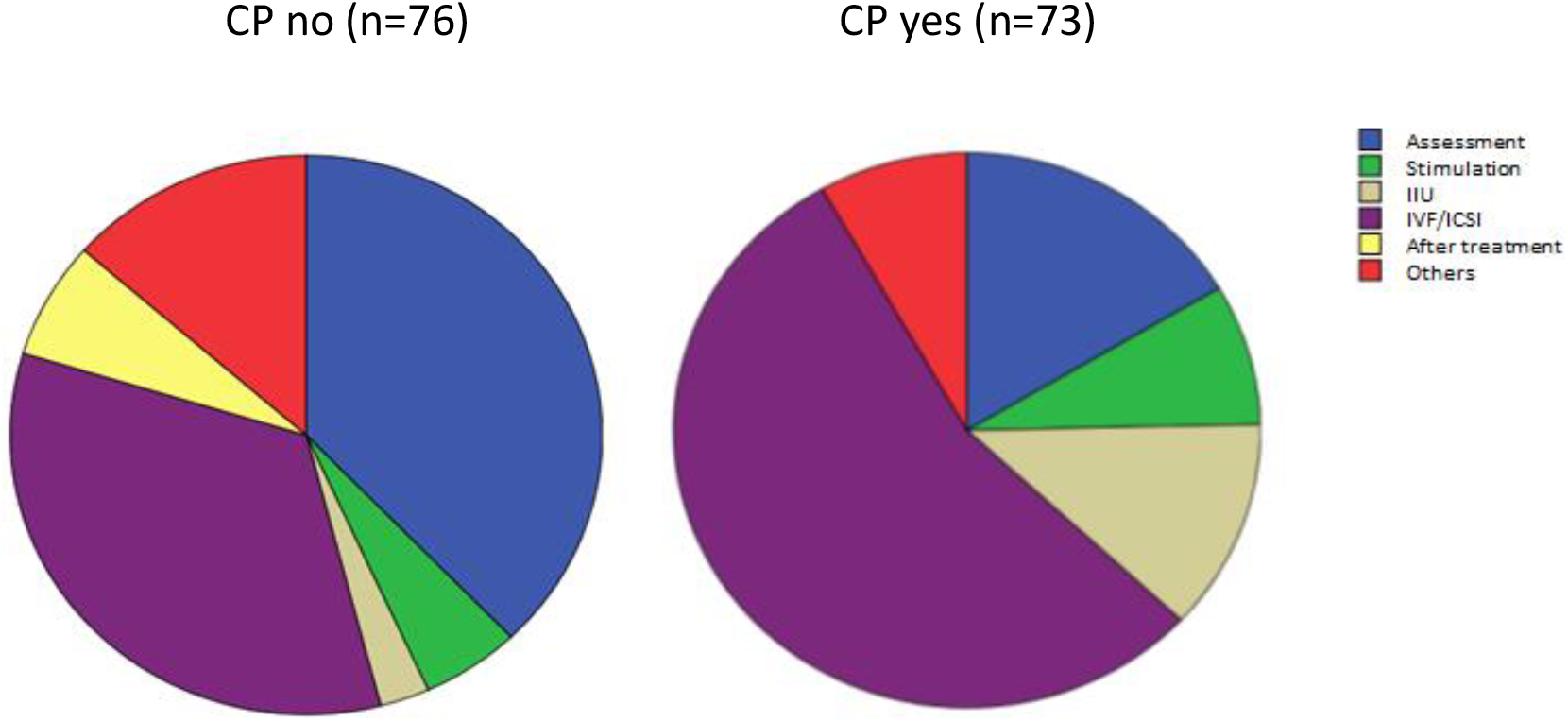
Sector diagram showing how the type of follow-up influencing the use of CP.

**Figure 3:**
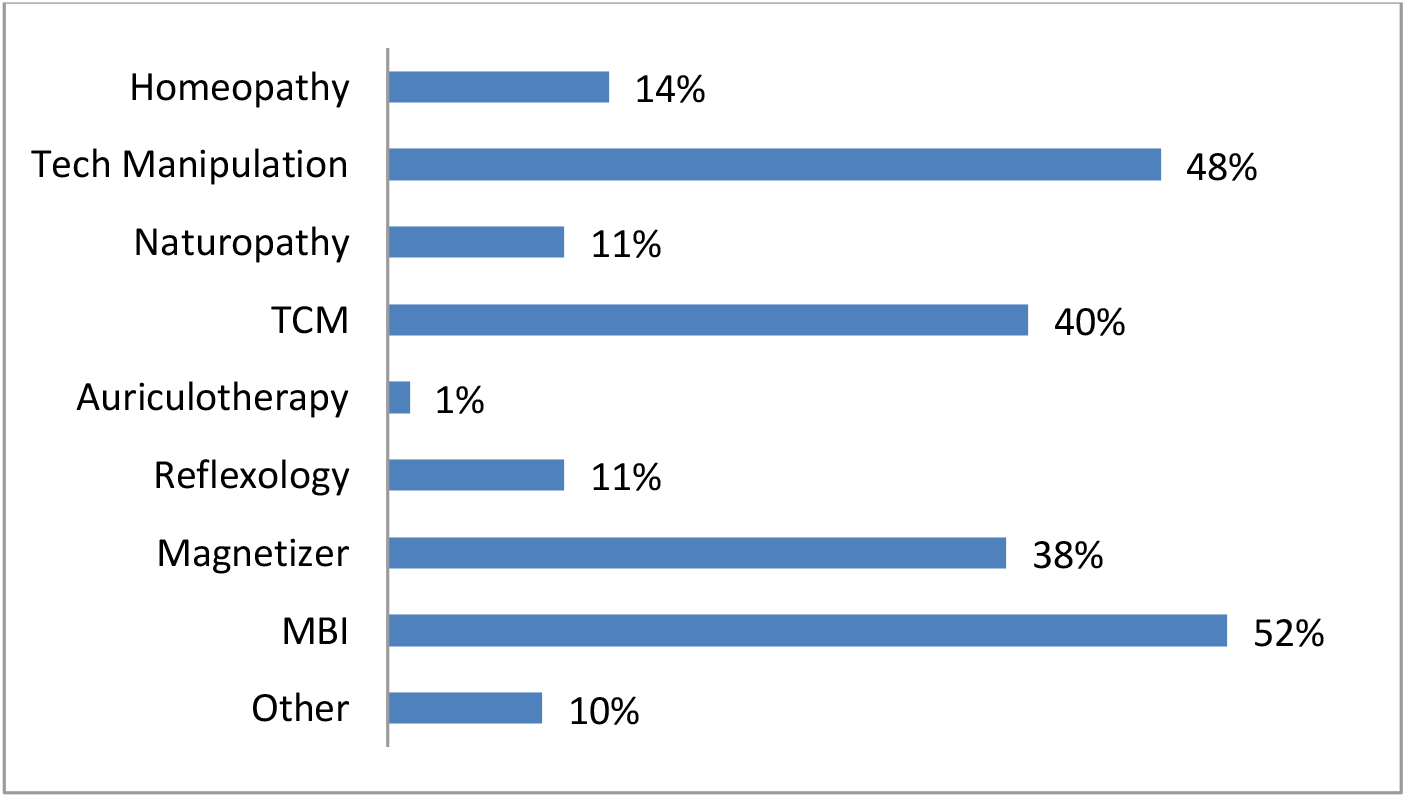
Type of CP used. Bar chart showing the percentage use of different types of CP in our population (n=73).

**Figure 4:**
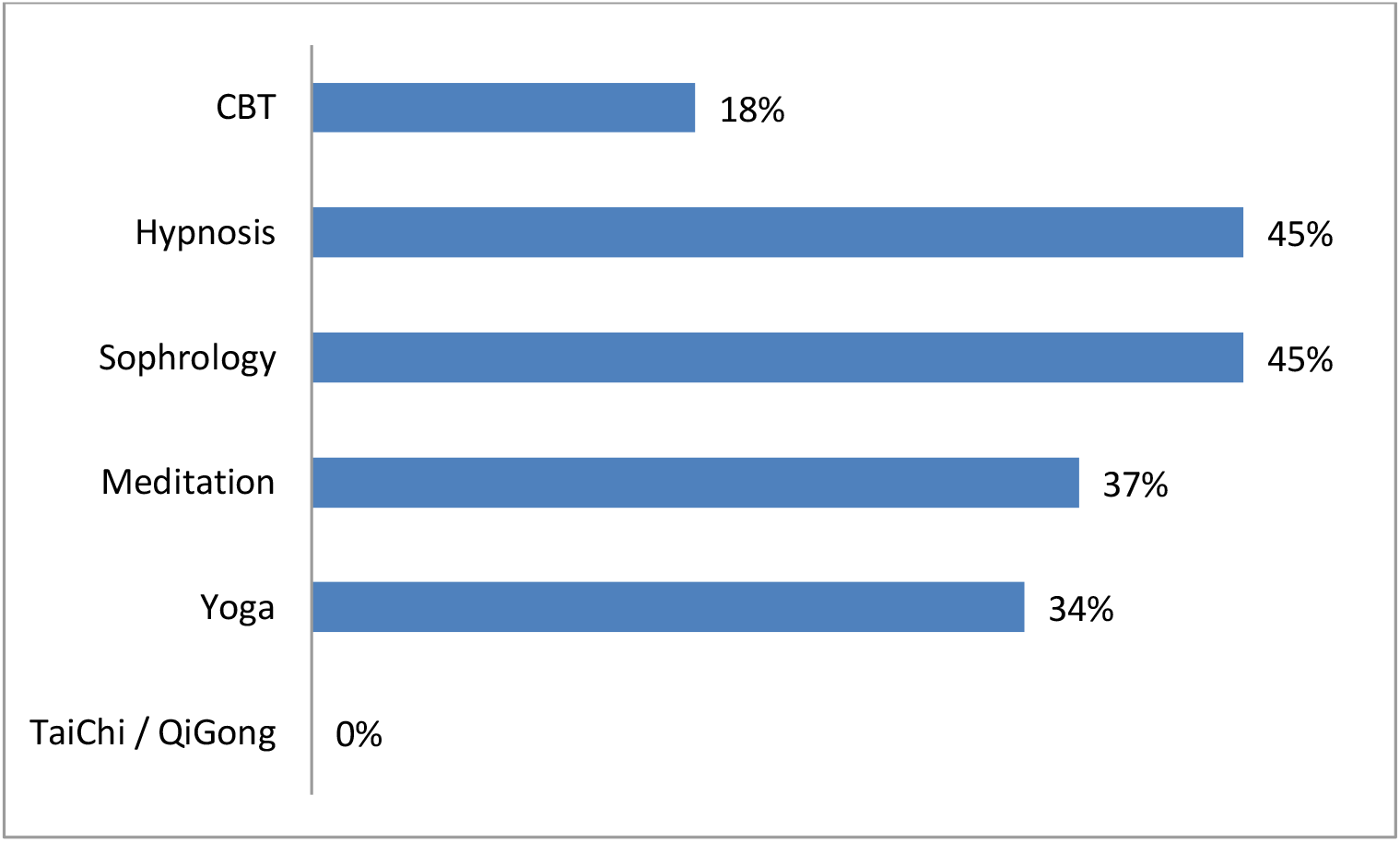
Bar chart showing the percentage use of different types of MBI in our population (n=38), CBT: Cognitive Behavioral Therapy.

### Role of Traditional Chinese Medicine (TCM)

TCM is a complete medical system that can have a positive impact on a woman’s journey through ART, not only by its effects on the gonadotropic axis function (8), but also by its potential effect on the biopsychosocial components associated with the diagnosis of infertility and its treatment. However, and to the best of our knowledge, there are no studies in the literature that looked at its global efficiency. Most studies have been centered around acupuncture or MBI, most probably due to methodological limitations, as well as the difficulties of finding licensed and experienced practitioners of TCM in Western countries.

### Acupuncture

Acupuncture is commonly used in the field of infertility, with the HEFA survey reporting a 26% utilization rate among women undergoing ART (11). Acupuncture seems to improve the well-being of patients by decreasing anxiety, and by reducing the pain felt during invasive procedures such as oocyte retrieval or embryo transfer (14). The first randomized controlled trials (RCT) on acupuncture in women undergoing infertility treatments are relatively recent. In 2002, a study including 160 women who did or did not undergo an acupuncture session, 25 minutes before and after the embryo transfer, showed a significant increase in pregnancy rates in the acupuncture group (42.5% vs 26.3%, P < 0.03) (15). Two other studies (16,17) also showed a significant improvement in pregnancy rates in IVF patients undergoing acupuncture on the day of embryo transfer. A 2006 study included 225 patients in two groups: the first group received acupuncture according to the TCM principles in the luteal phase, after the transfer, and the second group received a placebo (placebo points) (18). The pregnancy and implantation rates were significantly higher in the acupuncture group (29.4% vs 12.6% and 8.2% vs 3.2%, p<0.01, respectively). A recent study showed that, when compared with estrogen treatment, acupuncture and TCM significantly improved the endometrial thickness and the pregnancy rates in women with thin endometrium (19).

On the other hand, some studies reported contradictory outcomes and failed to show any positive impact of acupuncture. In a series of 150 patients, Domar et al. failed to replicate the results reported in the first 2002 study (20). Similarly, Smith et al. did not show any improvement in pregnancy rates in patients receiving a conventional acupuncture protocol when compared with patients receiving placebo (21). In a multicentric RCT in 2017, Fan et al. did not find any improvement in fertility in women with polycystic ovarian syndrome (PCOS) (22). Likewise, Madaschi et al. did not find any difference in the outcomes of 208 women undergoing IVF/ICSI between the group who had acupuncture and a control group (23) . Another 2013 RCT which included 62 women with PCOS undergoing IVF/ICSI showed a significant improvement of embryo quality following acupuncture, but there were no differences in the pregnancy rates after IVF compared with the control group (24).

Several systematic reviews and meta-analyses have attempted to synthesize the available data. While some failed to show any benefit (25,26), most of the recent ones, have reported a positive an impact of acupuncture on fertility and IVF outcomes (27–30).

In all, and based on the current available evidence, it is difficult to conclude on the impact of acupuncture on the pregnancy rates in ART. The discordance in the results is probably the consequence of the heterogeneity across the studies, most notably in the populations included, the indications, the experience of the practitioners, the choice of acupuncture points, and the whole course of treatment administered. Larger and better defined RCT, using more rigorous methodologies, are needed to confirm any potential positive impact of acupuncture.

### Mind-Body interventions (MBI)

Mind-body interventions are based on the interconnexion between the physical (bodily functions), psychosocial (emotions), and spiritual (meaning of life) well-being of a person, all of which play an important role when facing stressful life events.

ART are associated with several stressful factors that can significantly affect couples with infertility. This is why the “European Society of Human Reproduction and Embryology” (ESHRE) Psychology and Counselling Guideline Development” recommends psychosocial support as complementary therapy to infertility treatments (31).

Besides the discomfort it generates, stress is widely known to have deleterious effects on global health in general and on fertility in particular. Indeed, exposure to stress leads to the activation of certain areas of the brain, such as the prefrontal cortex, hippocampus, and amygdala. That signal is integrated by the hypothalamus which will in turn, via the central nervous system, stimulate the adrenal cortex to secrete adrenaline and noradrenaline and generate an adaptive response that impacts several systems in the body (cardiovascular, respiratory…). In parallel, the hypothalamus releases the corticotropin release hormone which triggers the production of adrenocorticotropin hormone (ACTH) by the pituitary, which will stimulate the production of corticosteroids. Corticosteroids can negatively impact the reproductive tract via several pathways: an interaction with the GnRH producing neurons, which can affect the gonadotropic axis and the production of FSH and LH, and an alteration of ovarian steroidogenesis and the development of the uterine mucosa (32).

The clinical studies on the impact of stress on reproduction have produced conflicting results. Some have failed to confirm a negative impact of stress on ART (33,34), while others have clearly demonstrated that negative effect (3,35). In a recent study, Haimovici et al. (36) found that an increase in the levels of stress-specific cytokines in the blood and the follicular fluid on the day of the oocyte retrieval was associated with decreased fertilization, implantation, pregnancy, and live birth rates (36).

MBI could modify the activity of the cerebral areas most impacted by stress (Prefrontal cortex, amygdala, hippocampus and hypothalamus). The use of the techniques has been linked to mood improvement and decrease of stress biomarkers. Furthermore, some of the MBI (Mindfulness, Yoga, Tai Chi and Qi Gong) could be able to affect gene expression, leading to an inhibition of the Kappa B pathway which is activated in case of chronic stress (37), as well as certain epigenetic mechanisms (38). Globally, MBI seem to be efficient in reducing stress during ART (32). Several meta-analyses have reported on the significant impact of MBI in reducing the psychological distress of patients undergoing ART, as well as improving the ART outcomes (39,40). A 2016 Cochrane review concluded that the effects of these interventions on pregnancy rates were uncertain due to the very low quality of the evidence, and that there was significant heterogeneity in the studies with regards to the patient population, the techniques and the protocols used (41). Other meta-analyses reported that these techniques had a significantly positive impact on the psychological health of patients without any improvement in the pregnancy and live births rates (42,43).

Globally, if the positive impact of MBI on ART outcomes (pregnancy and live birth rates) is yet to be definitely established, the literature is unanimous when it comes to their efficiency in improving the well-being of patients undergoing ART, and more specifically in significantly reducing the stress and anxiety levels.

The MBI used in the aforementioned studies were quite heterogenous: cognitive behavioral therapy, hypnosis, relaxation, respiration… (32,42,44). Only two studies, both by the same team, used a MBI approach based on TCM that included Tai-chi exercises, meditation and respiration (45). Both studies showed an improvement of the anxiety and well-being score of patients during ART, but did not show any improvement in pregnancy rates (46,47).

In our study, despite anxiety and fatigue being a major source of concern for patients, only 38 out of 150 resorted to MBI, and none used Tai-chi or Qi Gong (Figure 5). Tai Chi and Qi Gong combine slow movements and respiration exercises with relaxation and meditation, and are known to have a beneficial impact on the quality of life by reducing stress and anxiety levels, as well as relaxation and fatigue (48,49). These techniques could therefore play an important part in improving the well-being of women undergoing ART, and potentially improve the chances of success.

**Figure 5:**
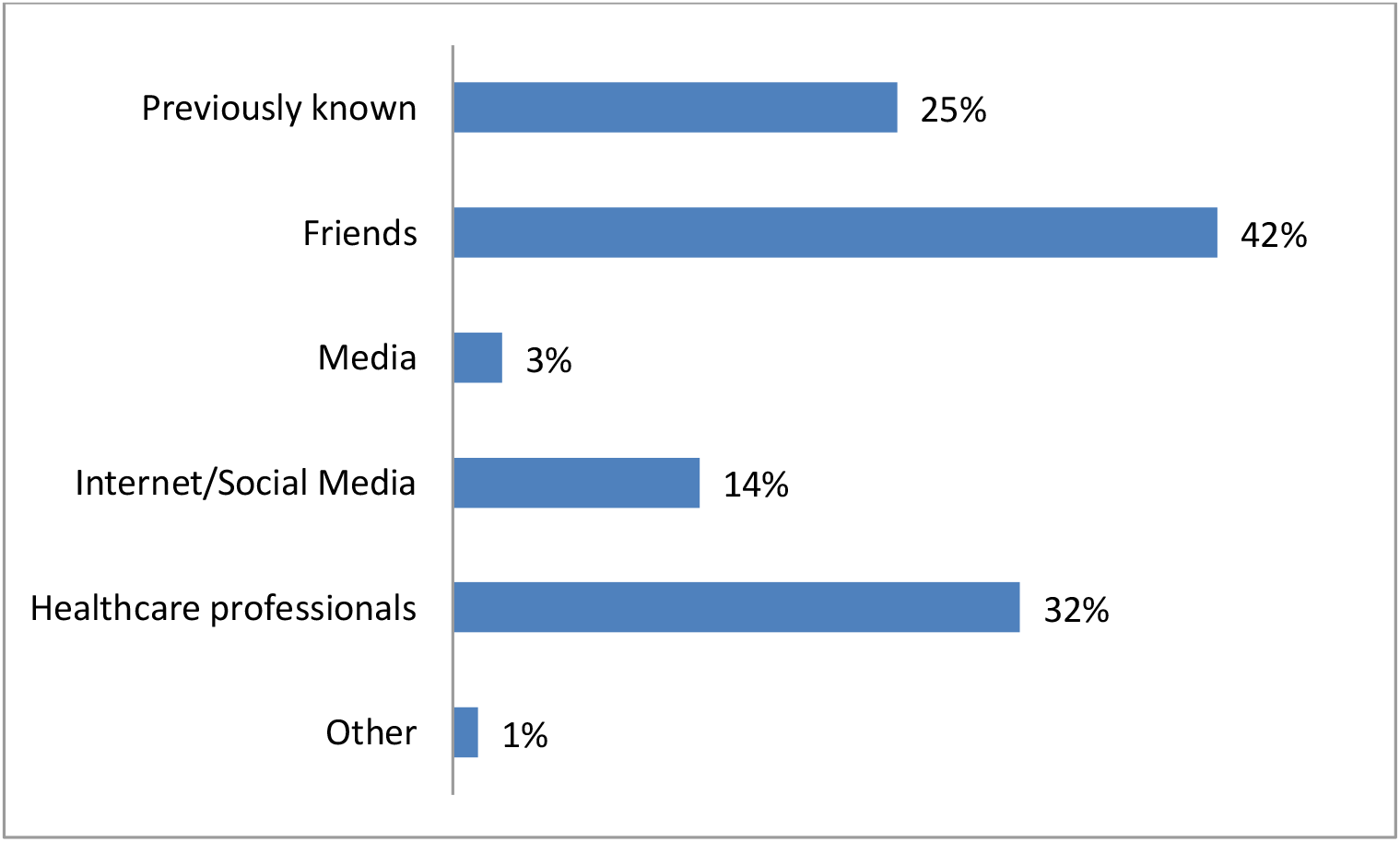
Bar chart showing the various means of CP information used by our patients.

**Figure 6:**
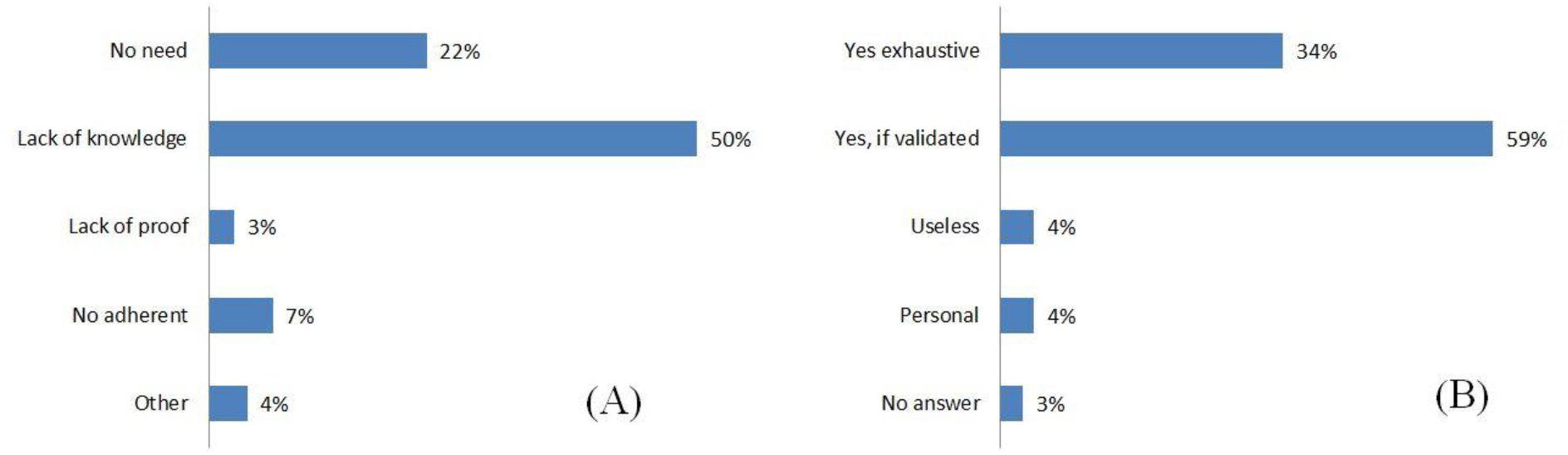
**(A)** Bar chart showing the main reasons why a part of our population doesn’t use CP (n=76). **(B)** Bar chart showing the desire of patients in our population to be informed about CP, whether or not they use them (n=150).

### Knowledge and information about Complementary Practices (CP)

In the current study, most women who resorted to CP were introduced to them by people from their personal network and by patients already using them. In 32% of cases, a healthcare professional was behind the introduction. It is worth noting that in a 2013 USA study, almost 90% of physicians interviewed reported being curious about complementary and alternative medicine, and 75% were interested in adding and integrating these practices to conventional medicine, as well as informing their patients about the methods available (14).

The lack of use of CP can be explained by the lack of knowledge and information. Indeed, among women who did not resort to CP, 50% were unaware of them, while a smaller percentage did not believe they needed them, and very few were doubtful and suspicious of their efficacy.

Out of the 150 women who answered whether or not they would like the physicians involved in ART to inform them about CP, only 12 thought that it was useless or too personal. The vast majority wanted their physicians to share global information about all these practices, and back it up with scientific proof.

For the physicians involved, the main problem is the large diversity of these methods, and the difficulty of knowing the benefits of each and every one. And even if for many of these techniques, there are several studies available in the literature, they are quite heterogenous and have lots of biases. However, newer and better studies are emerging and offering a better analysis of these practices. In particular, acupuncture and MBI seem to have a beneficial impact, at least in terms of quality of life and well-being of patients undergoing ART. The effect on pregnancy and live births rates remains under investigation.

## CONCLUSION

Our study has shown that women undergoing ART are interested in knowing about and resorting to complementary and alternative practices, with the aim of improving their chances of pregnancy as well as helping them deal with the stress and fatigue associated with ART. Among the different CP available, the ones based on body manipulation, acupuncture and mind-body interventions are the most coveted. Traditional Chinese Medicine includes all these techniques, and even though the jury is still out on its efficiency in improving pregnancy and live birth rates, information about its application and its benefits should be made available to all couples undergoing ART.

## Data Availability

All data produced in the present study are available upon reasonable request to the authors

## ACKNOWLEDGMENTS

We would like to thank all the people involved in the study. We also acknowledge the support of the «Institut National de la Santé et de la Recherche Médicale (INSERM) », of the « Centre National de la Recherche Scientifique (CNRS) », the Angers University, and the Angers university Hospital.

We thank the Integrated Centre of Chinese Medicine of the Sorbonne University for its expertise in TCM and related MBI

